# Preferences of health care workers using tongue swabs for tuberculosis diagnosis during COVID-19

**DOI:** 10.1101/2022.12.06.22283185

**Authors:** Renée Codsi, Nicole A. Errett, Angelique K. Luabeya, Mark Hatherill, Adrienne E. Shapiro, Katherine A. Lochner, Alexandria R. Vingino, Marlena J. Kohn, Gerard A. Cangelosi

## Abstract

Healthcare workers (HCW) who come into contact with tuberculosis (TB) patients are at elevated risk of TB infection and disease. The collection and handling of sputum samples for TB diagnosis poses exposure risks to HCW, particularly in settings where aerosol containment is limited. An alternative sample collection method, tongue swabbing, was designed to help mitigate this risk, and is under evaluation in multiple settings. This study assessed risk perceptions among South African HCW who used tongue swabbing in TB diagnostic research during the COVID-19 pandemic. We characterized their context-specific preferences as well as the facilitators and barriers of tongue swab use in clinical and community settings. Participants (n=18) were HCW with experience using experimental tongue swabbing methods at the South African Tuberculosis Vaccine Initiative (SATVI). We used key informant semi-structured interviews to assess attitudes toward two tongue swab strategies: Provider-collected swabbing (PS) and supervised self-swabbing (SSS). Responses from these interviews were analyzed by rapid qualitative analysis and thematic analysis methods. Facilitators included aversion to sputum (PS and SSS), perceived safety of the method (SSS), and educational resources to train patients (SSS). Barriers included cultural stigmas, as well as personal security and control of their work environment when collecting swabs in community settings. COVID-19 risk perception was a significant barrier to the PS method. Motivators for HCW use of tongue swabbing differed substantially by use case, and whether the HCW has the authority and agency to implement safety precautions in specific settings. These findings point to a need for contextually specific educational resources to enhance safety of and adherence to the SSS collection method.

## INTRODUCTION

TB disease, caused by *M. tuberculosis* (MTB), remains a major global cause of morbidity and mortality. Collection of the standard sample for TB diagnosis, sputum, presents safety risks to health personnel. Moreover, the process is difficult for many types of patients, and sputum testing is insensitive for certain types of TB. The availability of alternative, noninvasive sampling methods, which can easily be collected outside of the clinic, would improve worker safety, increase the efficiency of testing, and allow more active TB case finding in community settings.[1–3]

We and others have shown that MTB DNA is deposited on the oral epithelium during active TB, and can be detected by oral swab analysis (OSA).[4–15] In OSA, the dorsum of the tongue is brushed with a sterile swab. The swab head with collected material is deposited into a tube for MTB DNA detection by nucleic acid amplification testing (NAAT).[8] In this paper we focus on the two methods of sample collection using tongue swabs: HCW (provider)-collected swab (PS), or HCW-supervised self-swabbing (SSS).

Tongue swabs were developed in part to reduce the occupational health risks associated with sputum collection.[11] However, there may be new risks associated with tongue swabbing. In particular, the emergence of the SARS-CoV-2 virus created new potential threats to HCW collecting patient oral samples, and may have changed their attitudes toward such procedures.[16] In early evaluations of tongue swabbing, samples were collected by HCW or study personnel positioned in front of patients’ faces. This presented infectious disease exposure risks that were amplified by COVID-19. Therefore, a SSS approach, initially evaluated for COVID-19 tongue swab collection [6], was adapted for TB tongue swab collection.

Tongue swabbing for TB diagnosis remains investigational but is under evaluation in sites around the world.[11,17] Although qualitative assessments of preferences related to other aspects of TB diagnosis have been reported [18,19], there have been few such assessments of tongue swabs. Such studies are needed to understand the HCWs experience with tongue swabs in real world settings and how this impacts their willingness to use the new method for TB diagnosis. [20]

Implementation science is the scientific study of methods and strategies to increase the uptake of innovative, evidence-informed practice. A fundamental challenge of implementation science is identifying contextual determinants (e.g., barriers and facilitators) and determining which implementation strategies will address them.[21] Risk perception is a known determinant of implementation of evidence-informed practice.[19]

In the current study, we investigated HCW risk perceptions and their influence on HCW willingness to use PS and SSS for TB sampling, as compared to the gold standard sputum sample collection. We sought to characterize the facilitators and barriers of tongue swabbing for TB diagnosis during the era of COVID-19. We describe HCW willingness to use PS and SSS for TB diagnosis among HCW who make home visits and those who work strictly in the clinical setting in Western Cape, South Africa. Because tongue swabbing is still an experimental approach, we focused on South African workers who were among the few in the world to have used the method extensively prior to the study period.

## METHODS

### Strategy

For this study we used qualitative research methods, which deepen understanding by considering context and the complexity of multiple, overlapping, and sometimes conflicting themes.[22] Interviews and other qualitative research methods bring personal, social, and cultural knowledge into the research domain.[22]

### Data collection

We conducted and qualitatively analyzed key informant semi-structured interviews with HCW at the South African Tuberculosis Vaccine Initiative (SATVI) in Western Cape, South Africa, conducted between January 2021 and April 2021. Purposive sampling was used to identify HCW who met the inclusion criterion, namely experience working with both tongue swabbing and sputum collection for TB detection. Purposive sampling is a non-probability sampling technique frequently used in qualitative research to identify participants with lived experience or expertise that uniquely positions them to provide insights about a phenomenon of interest.[23]

Participants were referred to the study team by their site manager, and all referents who met the inclusion criterion were approached by the study team by email to invite them to participate. We continued recruitment until we observed saturation among meta themes, and additional interviews were determined to be unlikely to illuminate major new insights. [23–26]

An interview guide was developed *a priori* and reviewed by the study team and local investigators to ensure question clarity, local/cultural appropriateness, and robustness of the interview guide to address all study objectives. The interview guide was informed by the Extended Parallel Process Model (EPPM), [27] (**Figure 1**). EPPM is a risk communication theory that posits that individuals take action to control danger when they perceive that the severity and susceptibility are high and that they are competent to take mitigating action. We explored perceived self-efficacy (confidence in ability, knowledge, and skills to perform the method), perceived response efficacy (belief that the method works/works better than other methods), perceived threat susceptibility (belief that the HCW themselves are susceptible to TB/COVID-19), and perceived threat severity (belief that TB/COVID-19 is a formidable hazard). We explored how these constructs interacted with HCW willingness to perform specific sample collection methods in specific contexts.

**Figure 1:**
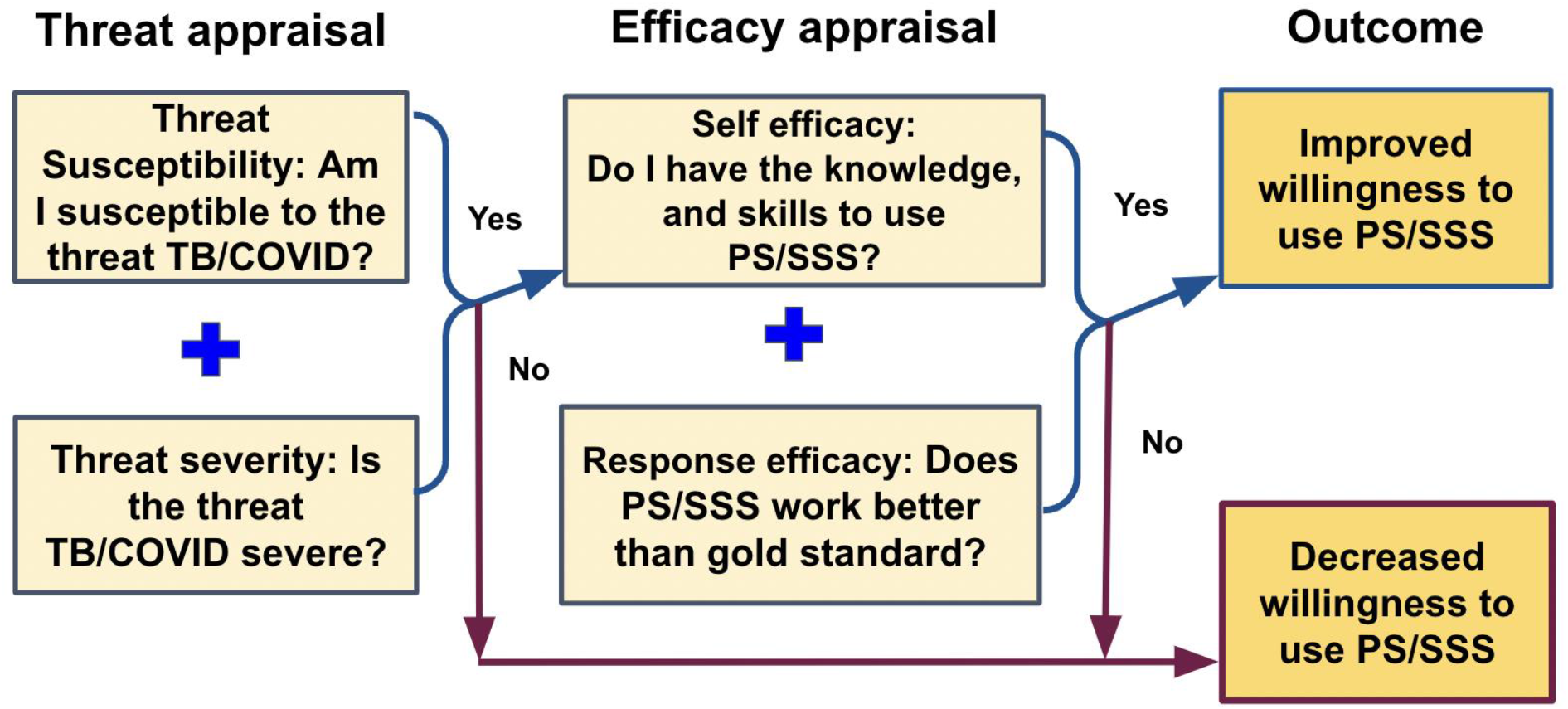
Theoretical framework for assessing preferences of health care workers using tongue swabs for TB diagnosis, using Witte’s Extended Parallel Process Model.

All interviews were audio recorded. Because travel was restricted during the COVID-19 pandemic, interviews were conducted remotely over Zoom in English. Interview audio files were professionally transcribed by TranscribeMe! (San Francisco, CA) and reviewed for accuracy.

### Data analysis

We used a two-phase approach to analysis. First, we adapted Hamilton’s rapid qualitative analysis methods to summarize key findings by research objective.[28] Interview transcripts were read and re-read by the primary interviewer (RC). An interview contact sheet was created for each interview, summarizing key takeaways related to each of the study objectives. These contact sheets were shared back with participants for confirmation of accuracy. This process, known as member checking, provides an opportunity to enhance qualitative credibility.[29] We then entered data from each of the interview contact sheets into a matrix. The matrix was used to synthesize key takeaways across interviews to identify commonalities and counterpoints.

Next, transcripts were coded and thematically analyzed. Deductive codes were developed based on the interview guide and underlying theory (EPPM) and research objectives. For the inductive phase, we re-read interview transcripts and member checking summaries to identify emergent themes. These were formalized into codes and given a definition based on our evolving understanding of the theme. Codes, their definitions, and examples of when to apply them were formalized into a codebook.

Codes were applied to transcript data using NVivo version 12 for Mac qualitative data analysis software. Approximately 20% of data (4 transcripts) were co-coded by a second researcher to confirm face validity of code construct definitions and to enhance the reliability of the codebook. Coding discrepancies were reviewed and discussed to identify opportunities for code definition or codebook revision, and resolved by consensus. A single coder (RC) used the updated codebook to code the remainder of the transcripts. The interview guide is provided in Supportive Information.

A code network map was used to align codes with each of the research objectives.[30] Data associated with each relevant code was synthesized and summarized to identify key themes and counterpoints by research objective.

### Ethics

The University of Cape Town Institutional Review Board (IRB00001938) and the University of Washington Human Subjects Division (STUDY00011556) reviewed and approved this study. All participants provided written informed consent.

## RESULTS

### Participant characteristics

Participants had from 5 to 20 years of experience collecting samples for TB diagnosis. HCW participant demographic information is outlined in **Table 1**. A majority (15/18) make both home visits and work in a clinic, while a few (3/18) work strictly in the clinic setting. Six participants work in the same community where they live. Most of the participants (16/18) work with patients who are between the ages of 18 and 65 years. One participant sees patients who are between 5 and 18 years old, and 2 participants see patients who are less than five years old. A majority (15/18) speak more than one language at work.

**Table 1:** Demographic data of key informants.

| **Participant Demographics (N=18)** | **N (%)** |
| --- | --- |
| Female | 14 (78) |
| Male | 4 (22) |
| Works with patients age 0-5 (years) | 2 (11) |
| Works with patients age 5-18 (years) | 1 (6) |
| Works with patients age 18-65 (years) | 16 (89) |
| Works strictly in clinics | 3 (17) |
| Makes home visits | 15 (83) |
| Lives and works in the same community | 6 (33) |
| Speaks more than one language at work | 14 (78) |
| Previous TB disease diagnosis as of April 2021; reported as occupationally acquired | 2 (11) |
| Previous COVID-19 diagnosis as of April 2021; reported as community acquired | 3 (17) |

Participants were also asked about their health history related to TB and COVID-19. Two of the 18 participants had a previous TB diagnosis and treatment. Both cases were reported as due to occupational exposure. Three of the 18 participants had a previous COVID-19 diagnosis, with all three cases reported as community exposure.

### Preferences

Overall, the majority (15/18) of HCWs preferred tongue swabbing of any kind over sputum sample collection method. PS was the most preferred (9/15). Only 3 respondents preferred sputum sampling over tongue swabbing. Motivators of swabbing differed substantially by use case, and whether the HCW has the authority and agency to implement safety precautions in specific settings. Below, we discuss key facilitators and barriers to PS and SSS use (**Figure 2**).

**Figure 2.**
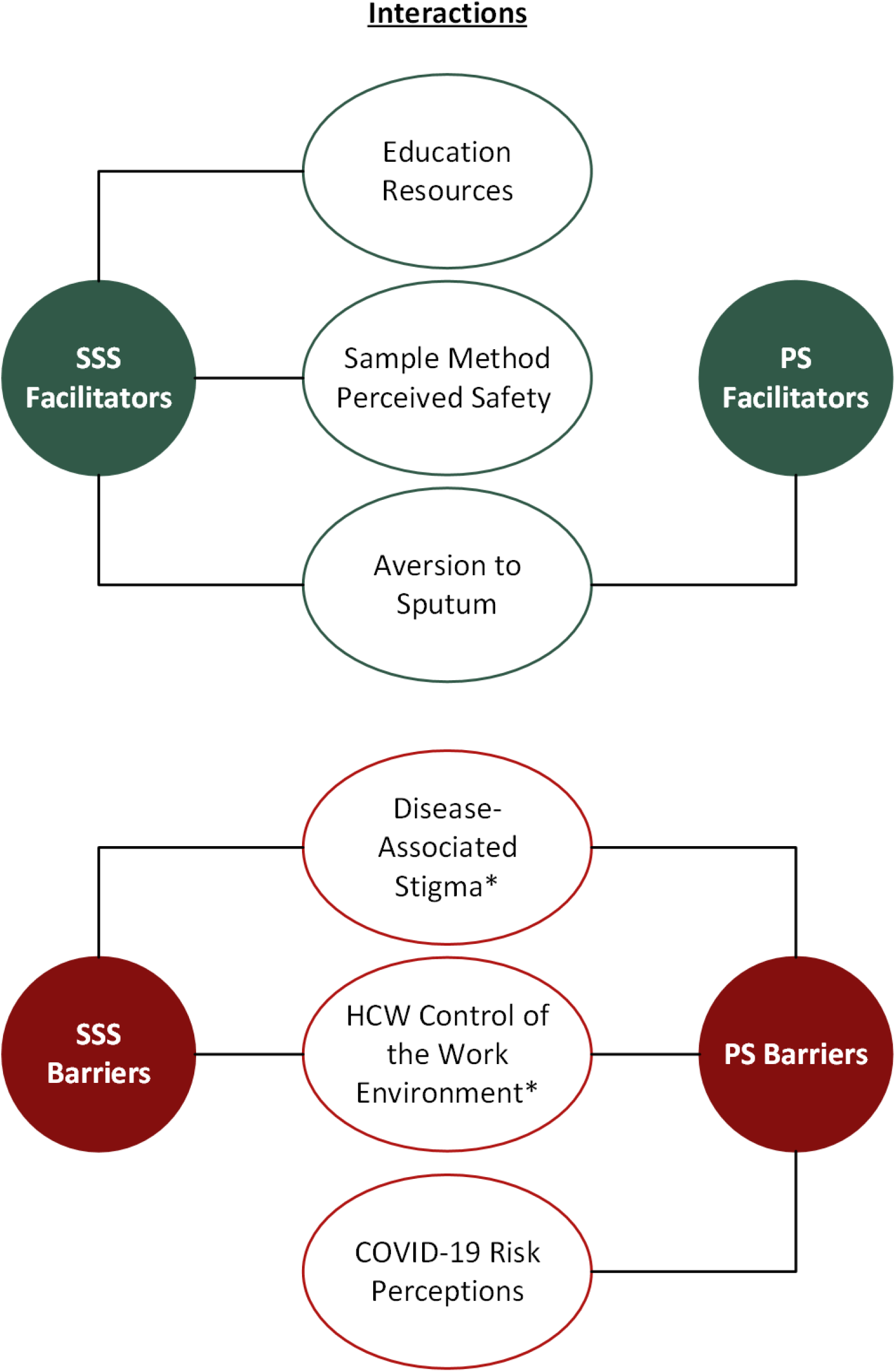
Key theme interactions acting as facilitators and barriers to PS/SSS sampling methods. * Themes related to home visits only; Disease associated stigma and HCW control of the work environment.

## Facilitators

### Aversion to sputum (PS and SSS)

Almost all HCWs interviewed shared that they do not like looking at and collecting sputum samples. HCWs must confirm the color and volume, which was reported to be an unenjoyable process. They also shared that sputum can be messy if it is not collected properly and if the jar is not closed properly. As one HCW who works in the community and the clinic shared, “*Sometimes the participants don’t close the lid of the stuff on the sputum jars and then when you get there, everything is out of the sputum jar. The swab is less messy than sputum and you know that you get a sample that’s not going to leak or something like that*.*”*

HCWs who work with infants and children expressed population-specific challenges with obtaining sputum. Standard methods for children include invasive and poorly tolerated procedures such as gastric lavage. HCW expressed the need for another sampling method and that they were interested in trying the PS method with infants and children. As one who works with children in the clinic shared, *“Inducing sputum production in kids is difficult for parents to see their child go through and there is loss to follow up because parents don’t want to put their kids through that. It would be interesting to see if we can start to use swabs on children*.*”*

However, one HCW shared concerns about collecting PS from infants, as the infants need to be still during the sample collection process.

### Perceived safety of the method (SSS)

All participants expressed that they have the knowledge to protect themselves from the risk of TB, and most also shared that they have the knowledge and skills to prevent COVID-19 exposure at work. The majority of HCW interviewed identified SSS as their preferred method of sample collection compared to both PS and sputum sampling due to perceived safety. They reported perceptions of being able to keep a safe distance while observing the patient to make sure they were performing the sample collection correctly. correctly. As one HCW who both worked in the community and clinic shared,*”SSS is safer for the family members in the house and if it is done at the clinic then it is safer for the other staff members too*.*”* Another, who also worked in both contexts, revealed similar sentiments, *“I prefer SSS for anyone that is not a child or elderly because they* (children and elders*) will not be able to listen and do it right*.*”*

The majority of the HCW shared that being close to the patient in front of their mouth while using the PS method made them hesitant, especially with the dual threat of TB and COVID-19 exposure. On the other hand, some HCW who worked in both the community and clinic shared that they preferred PS due to perceptions that patients, particularly intoxicated patients, would not perform SSS correctly: *“I prefer taking the risks that come with OSA* (PS) *to supervised self-swabbing. I do not have confidence that patients can accurately perform* (SSS).*”* Another, who also worked in both contexts, shared, *“I prefer* (SSS) *but only works for those who are not drunk and can follow directions*.”

### Educational resources to train patients (SSS)

All of the participants advocated for culturally appropriate training modules to help their patients learn how to effectively self-collect the samples (SSS). Several participants shared concerns that they would have to resort to the PS method in the absence of useful educational materials. The potential availability of educational materials is a key facilitator increasing their willingness to use SSS in particular, and by extension, tongue swabbing in general.

Not all HCWs expressed confidence that their patients could follow directions to collect the sample safely and effectively themselves. Participants who make home visits expressed concerns regarding certain patients who present the greatest challenges such as patients who are intoxicated, children, elderly, and patients who are too sick to follow directions, or settings where they have limited control of the working environment. The majority of the HCWs shared that with improved educational materials and resources for themselves and their patients, they can envision SSS being implemented with their patients with the cognitive abilities to follow directions.

Overall, HCWs shared that they have comprehensive training for sputum and PS but that they would like training on SSS particularly in response to the threats of COVID-19. Many HCW expressed that they would like to be trained on how to facilitate SSS; they also shared ideas for improving training using videos, animations, and photos to guide patients through SSS regardless of literacy level or language spoken. As one HCW who conducts home visits and works in the clinic shared, *“I need to have videos. Nowadays, people are very lazy to read. So many people*…*are now very digitized to their phones. So maybe a free website that will give some free education pertaining to certain signs and symptoms and diseases*.*”* Another, who also works in both contexts, went on to say, “*Cartoons would be great to help guide the patient through swabbing with an explanation, then patients could successfully self-swab*.*”*

HCWs emphasized the need for training to be accessible to patients, acknowledging both technology access realities and language needs. Some shared that videos could be played in a clinic waiting room, but the expectation should be that all training be conducted at the clinic as many patients lack smartphones or data necessary to stream the videos. One HCW shared that training materials, including videos, should be available in English, Afrikaans, and Xhosa.

## Barriers

### Disease associated stigmas

HCWs discussed how the HIV-TB co-epidemic in South Africa affects TB stigma and their ability to manage risk. The majority of HCW making home visits described the importance of protecting patient confidentiality and preventing stigma associated with TB testing and diagnosis. As one HCW who works in both the community and clinic shared, *“Most of the people here in our community, they think when you have TB, they also have HIV. So some of them, they are afraid to just confide in their family*.*”*

HCWs described how this desire to maintain confidentiality and prevent stigma limited their ability to enforce social distancing and other infection control measures in home visit settings; it also made home-visit HCWs reluctant to use tongue swabbing because of increased exposure and infection risk for PS/SSS sample collection without distancing or infection control measures.

In addition to disease associated stigma with HIV, there were separate concerns about oral hygiene. We have reported that OSA signals are somewhat stronger in samples collected early in the day, relative to samples collected later in the day.[9] Therefore, in some OSA research protocols, samples are collected in the early morning and patients are asked not to brush their teeth before sample collection. HCWs expressed concerns with the logistical challenges with early morning sample collection before the patient eats, brushes their teeth, or goes to work.

HCWs also shared their perceptions of patient comfort when taking early morning tongue swab samples, stating that patients may be shy and concerned with their early morning breath. HCW shared that collecting samples during the daytime and not before brushing their teeth in the morning may reduce the barrier that the patients feel when opening their mouth before they get a chance to brush their teeth so early in the morning.

### Control of the work environment in community settings

HCW in clinic settings reported that they had control of their environment to implement training to mitigate risk while collecting samples, were treated professionally by patients, and had authority to direct patient behavior in the clinic. However, HCW in home visit settings shared that they did not have the same control of their environment and often did not have the authority to control their patients’ actions especially when related to others in the household. As one who worked in both contexts shared, “*If you’re at a patient’s home, their kids, they will come and touch you. So, we are in high risk at the home of the participant*.”

Female HCW expressed concern with being treated more like an “auntie” (a family friend who doesn’t challenge the norms of the household) than as a professional. HCWs expressed greatest concern with being able to keep their distance and abide by safety standards when in the home. The majority of the participants who make home visits shared examples of situations when they did not have the authority or agency to protect themselves, which impacts their agency to respond to perceived threats; this was exacerbated by cultural stigma, patient discomfort with sample collection in the early morning, and the context of COVID-19. HCW struggled to balance collecting the samples safely to prevent TB or COVID-19 transmission and being culturally respectful when entering someone else’s home. One HCW shared that they wanted uniforms for their work that were washed at work and not taken home, outdoor facilities for patients to produce sputum samples, and larger clinic spaces to accommodate social distancing.

### COVID-19 risk perception

Although all HCWs stated that sputum collection presented challenges and that tongue swabbing would be preferred, they expressed a variety of barriers to tongue swabbing related to their perceived risk of COVID-19 while collecting samples. More than half of the HCW expressed that the threat of COVID-19 exposure impacted their willingness to stand in front of the patient to collect the samples (PS). Some HCW reported that this changed their preference to SSS. For instance, one HCW who works only in the clinic shared, *“Self-swabbing with me there guiding at a distance would be better now with COVID*.*”* Another who works both in the community and clinic shared similar sentiments, *“Before COVID I preferred to do the tongue swab but now with COVID I would prefer to do the SSS. It (SSS) is also safer for the family members in the house… if it is done at the clinic then it is safer for the other staff members like cleaners, security and register*.*”*

However, two HCWs who previously preferred PS reported that they preferred sputum sampling over any self-swabbing methods in the context of COVID-19. As one HCW who worked both in the community and the clinic shared, *“For now, because of the pandemic, I prefer the sputum over the (provider) swab. I don’t want to be so close. But before the pandemic, the (provider) swab was a good alternative to sputum*.*”*

A few participants described how they perceived that fear of COVID-19 and community and occupation controls worked to reduce the spread of TB at work and in their community. The COVID-19 pandemic has introduced new trainings to the curriculum such as social distancing and reinforcement of use of PPE. During COVID-19, PPE has also become more readily available, and adherence to PPE protocols has become stricter. Precautionary behaviors for TB have had higher adherence due to fear of contracting COVID-19. In addition, the novelty of the hazard of COVID-19 influenced their risk perceptions. HCW were more concerned with contracting COVID-19 than TB at work. As one HCW who worked exclusively in the clinic shared, *“Before COVID I got TB because a patient coughed in my face in the clinic. I was not wearing a mask because I did not know he had TB. That would not have happened now because I am afraid of COVID and wear PPE all the time at work*.*”*

In addition to COVID-19, HCWs described other perceived threats to personal safety. Among HCWs doing work in the community, as opposed to a clinic setting, threats to personal security were a common theme for HCWs in home visit settings. While this came up in the context of this study on tongue swab barriers and facilitators, it is not specific to OSA and could apply to any work conducted by the HCWs in community and home settings for any health conditions. HCWs described threats to their personal security as a prohibitive barrier to performing all sampling methods (sputum collection, PS, and SSS), and was noted by both male and female HCWs. Both logistical and socioeconomic factors influenced personal security while doing their work. Eight of 18 HCWs shared examples of risks to personal security. One who works in the community shared a past experience, *“As I was looking, this other guy was carrying a knife. I did not feel safe going out of the car to walk up the stairs to the participant’s home*.*”*

## DISCUSSION

This study explored HCW willingness to use tongue swabs, especially with regard to perceptions of risk of occupational exposure to TB and COVID-19. Some facilitators and barriers reported by HCWs were germane to tongue swabbing in general, while others were specific to different methods (PS or SSS). For both PS and SSS, aversion to sputum was reported to support HCW willingness to use the methods. For SSS specifically, perceived safety of the sampling approach, and educational resources to train patients how to collect SSS samples were also described to positively influence HCW willingness to use tongue swabbing. Barriers to both PS and SSS included disease-associated stigmas with TB infection, and HCW control of their work environment when making home visits. For PS specifically, COVID-19 risk perception was identified as an additional barrier. Motivating reasons behind HCW willingness to use tongue swabbing varied substantially by HCW environment (clinic versus community settings), particularly in community settings where HCW perceptions of their authority and agency to implement safety precautions at home varied by case setting. These issues may require contextually specific solutions.

Collecting samples for TB diagnosis poses risk to HCWs, particularly in areas where resources for aerosol containment from sputum sampling methods are scarce. Notably, while HCWs reported high levels of self-efficacy to perform PS and SSS, some reported that their perception of patient self-efficacy for the SSS method informed their preference for PS, despite perceptions that SSS was safer for the HCWs themselves. Perceived patient self-efficacy has been reported to influence HCW willingness to use other types of diagnostics or put themselves at risk.[31] Accordingly, HCWs pointed to the need for educational materials for patients, including illustrations and videos, to improve their capability to perform SSS correctly. This solution has the potential to both improve implementation of SSS and support HCW willingness to perform a safer and preferred method of sample collection (PS). As described by our study participants, these materials should be made in multiple languages and should be available in the clinic, as the patient population of interest may lack the technology and resources to review them beforehand. HCW could be provided with digital educational materials (and airtime/data) to show participants in their homes. Although this was not specifically suggested by HCW, it would meet the criteria HCW specified of being accessible to patients, and would allow viewing in the home health community setting without relying on clinic resources.

HCWs making home visits pointed to the need to protect patient confidentiality and prevent stigma associated with TB testing and diagnosis. HCWs described how this need limited their ability to enforce social distancing and other infection control measures in home visit settings. In addition to culture stigma associated with HIV, there were separate concerns about oral hygiene. We reported recently that oral hygiene does not strongly affect OSA (tongue swabbing) results.[32] This finding may help mitigate this barrier as updated collection protocols may not need to require that HCW collect swabs first thing in the morning or before participants brushing teeth. The flexibility for patients to brush teeth and provide samples at any time of day may help to increase acceptance of tongue swabs.

Our study had limitations. It was conducted in a single site in South Africa, which may limit the generalizability. However, as noted above, this deep context-specific inquiry showcased the importance of local context in the determination of facilitators and barriers to HCW willingness to use tongue swabbing. Moreover, as participants were sampled from one of the first sites with considerable experience using this approach, this allowed for early formative research on implementation. Additional site-specific qualitative inquiries are thus recommended.

Qualitative interviews were performed over Zoom by a U.S.-based researcher (RC) in participants’ non-primary language, introducing opportunities for misinterpretation of both questions and answers. To promote qualitative trustworthiness, we introduced a number of credibility checks. First, our study team included in-country collaborators, who reviewed our study protocols and interview guide to ensure question clarity and local and cultural appropriateness. Interviews were recorded and professionally transcribed, and professional transcriptions were reviewed by the research team for quality assurance, to ensure robust data capture. We also employed member checking [29], wherein we provided interviewees with a summary of our key takeaways from their interview, aligned with our research objectives.

Participants were offered the opportunity to review and provide feedback on this interpretation. In conclusion, interviews with 18 HCWs in South Africa supported the acceptability of PS/SSS as a promising method for collecting TB diagnostic samples from patients while reducing occupational risks. Interviewed HCWs preferred PS/SSS over sputum sampling, attributed to their aversion to sputum and perceived relative safety of tongue swabbing, with SSS being perceived as the safer option. However, some HCWs described that they were deterred from using the preferred method (SSS) because of their lack of confidence in patients’ ability to collect the sample correctly. Accessible educational resources, both in the terms of language and technology access, targeted at patients could thus improve SSS uptake and HCW occupational health and safety. Disease-associated stigma, lack of control over the workplace environment, and COVID-19 risk were also identified as barriers to tongue swab sampling. These findings support the need for formative, context-specific implementation research in parallel with the rollout of novel diagnostic sampling approaches.

## Data Availability

Data including code book, rapid analysis framework matrix, SOP's for supervised self-swabbing and provider swabbing, and supportive quotes for barriers and facilitators, are available upon written request.

## ACKNOWLEDGMENTS

We are grateful to the HCWs who agreed to be interviewed for this study as well as the patients, communities and clinical teams in Western Cape for their longstanding participation in TB research. We are specifically grateful for the contributions of Danelle Van As, SATVI study coordinator and research nurse who was essential for data collection, site management and operations which facilitated this work. This work was supported by the Bill and Melinda Gates Foundation (INV-004527, OPP 1213054), and by NIH grant U54EB027049.

